# Disease and Economic Burden of Stillbirths in India in 2019

**DOI:** 10.1101/2023.10.16.23297071

**Authors:** Vidhi Wadhwani, Divya Shrinivas, Sweta Dubey, Siddhesh Zadey

## Abstract

**Background:** India has the highest number of stillbirths worldwide. However, disease and economic burdens assessment of stillbirths in India remain missing.

**Methods:** We conducted a retrospective analysis using stillbirth data from the health management information system (HMIS), and civil and sample registration systems (CRS and SRS) for India and its 21 states. We calculated disease burden in terms of disability-adjusted life years (DALYs) and economic burden in value of life-years (VLYs). We conducted multiple sensitivity analyses for DALY and VLY assessments.

**Results:** Indian HMIS reported 263,342 stillbirths in 2019. Stillbirths led to 18.3 million DALYs and a monetary loss of INR 6.4 trillion. Five states contributed to more than 40% burden. Sensitivity analyses showed consistency in findings.

**Conclusion:** Due to their disease and economic burdens, stillbirths should be prioritized in the public health agenda.

## Introduction

The World Health Organization (WHO) defines a stillborn as a baby who dies after 28 weeks of pregnancy but before or during birth.[1] In several countries, stillbirths equal neonatal mortality in absolute count. The majority of stillbirths can be averted by improving family planning, antenatal, and obstetric care. While pediatric disease burden is high on the global agenda, stillbirths are often neglected. One depiction of neglect is the absence of stillbirths in the Global Burden of Disease (GBD) study estimates.[2]

In part, the neglect is due to methodological challenges in ideating and estimating the disability-adjusted life-years (DALYs) for a condition that involves “pre-birth” mortality. DALYs are calculated as a sum of Years of Life Lost due to prematurity (YLLs) and Years Lived with Disability (YLDs).[3] Since YLDs are zero for stillbirths, DALYs equate to YLLs. Investigating stillbirth disease burden beyond mortality can help assess the overall toll of the problem. It also ensures the inclusion of stillbirth in the disease burden discourse, which is growing in popularity for policymaking and health investments.

Investments in health often have to rely on evidence that depicts the disease’s economic burden. There are multiple ways to capture such a burden. Previously, cost-of-illness assessments that investigate direct and indirect costs have noted that stillbirths can cost families anywhere between 10% and 70% greater than a live birth.[4] While the literature is sparse, it is argued that the economic burden of stillbirths is often underestimated, often neglecting the emotional, psychological, and social burden that comes with it.[5] Another approach to comprehensively assess economic burden is the value of life-year (VLY) or full-income approach.[6] The VLY approach uses improvements in gross domestic product (GDP) and life expectancy to depict the overall impact of healthcare improvements on populations. For a South Asian country, it has been estimated that the value of a life-year is 2.8 times the GDP per capita.[6]

As per the 2020 United Nations report, India contributes to 18% of global stillbirth mortality and has the highest absolute number of such deaths of any country.[7] In response, the country has launched the Indian Newborn Action Plan (INAP), which aims to reduce the stillbirth rate to a single-digit value by 2030.[8] However, INAP does not look into the stillbirth burden, and there are no such assessments for India and its states.

Our work had two main aims. First, we estimated the disease burden of stillbirths. We used the YLL approach as our primary analysis, and an approach proposed by Kant for sensitivity analysis.[9] Second, we estimated the economic burden using the VLY approach.

## Methods

We calculated the disease burden in terms of disability adjusted life-years (DALYs). DALYs are calculated as the sum of years of life lost due to premature mortality (YLLs) and years of healthy life lost due to disability (YLDs).[3] YLDs are zero for stillbirths, so DALYs are equal to YLLs. Here, YLLs are calculated as the product of life expectancy and the number of stillbirths **(Equation 1)**.

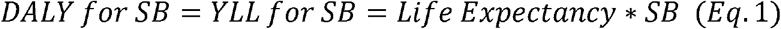

For our primary analysis, we extracted values for live births and stillbirths from the Health Management Information System (HMIS) and life expectancy data from the sample registration system (SRS) for India and its 21 states and UTs for the financial year 2019-20.[2,10] For comparison across data sources, we calculated DALYs using stillbirths derived from SRS (using still total birth rate data) and civil registration system (CRS) for India and its 21 states/UTs.[10,11]

For sensitivity analysis, we calculated the disease burden of stillbirths via a framework proposed by Kant. This framework defines stillbirth mortality in line with neonatal mortality, i.e., using live births instead of total births (i.e. stillbirths and live births) as the denominator. This still-live birth rate adjusts the population-level life expectancy for stillbirths. The derived life expectancy was used to calculate DALYs(see **Web Annexure 1**).[9] Similar to our primary analysis, we calculated DALYs using all three data sources (i.e. HMIS, SRS, and CRS).

We compared the DALY values obtained from the primary and sensitivity analyses using percentage differences for India and its 21 states /UTs **(Equation 2)**.

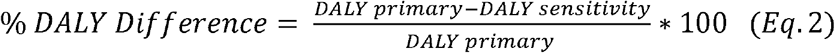

Further, we used non-parametric Spearman’s correlation coefficient to establish an agreement between the two analyses. We conducted these analyses only for HMIS data.

We calculated the economic burden using the value of life-years (VLYs) approach, which assigns a monetary value to each life-year (see **Equation 3**). VLYs consider the gains in GDP and population-level life expectancy in the past decades due to public health interventions. The Lancet Commission on Investing in Health has noted that the levels of gains vary across World Bank regions.[6] For South Asia, the value of one life-year is equivalent to 2.8 times the per capita GDP at a 3% discount rate.

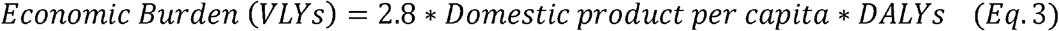

We calculated three VLY values for India and its 21 states and UTs using per capita GDP values. For comparison across data sources, we also used per capita net domestic product (NDP) and non-health gross domestic product (NHGDP). NDP signifies the net value of all goods and services produced in a region and NHGDP is the per capita GDP minus the per capita health expenditure of the region, which is the most conservative measure.

We extracted the GDP and NDP values from the Reserve Bank of India (RBI) Handbook of Indian Statistics 2019-20 and per capita health expenditure values from National Health Accounts (NHA) for the financial year 2019-20.[12,13] All monetary values were expressed in Indian National Rupees (INR) for the year 2019 and were also converted to USD based on an exchange rate of 1 USD = 70.394 INR.[14]

## Results

In 2019, HMIS reported 263,342 stillbirths in India. Uttar Pradesh, the most populous state, had the most stillbirths (45,556) while Lakshadweep had the least (5). Nationally, stillbirths led to 18.35 million years of life lost. Across states/UTs, Uttar Pradesh had the highest disease burden (2.99 million life-years) while Himachal Pradesh had the lowest (72,000 life-years) **(Figure 1a)**.

**Figure 1a:**
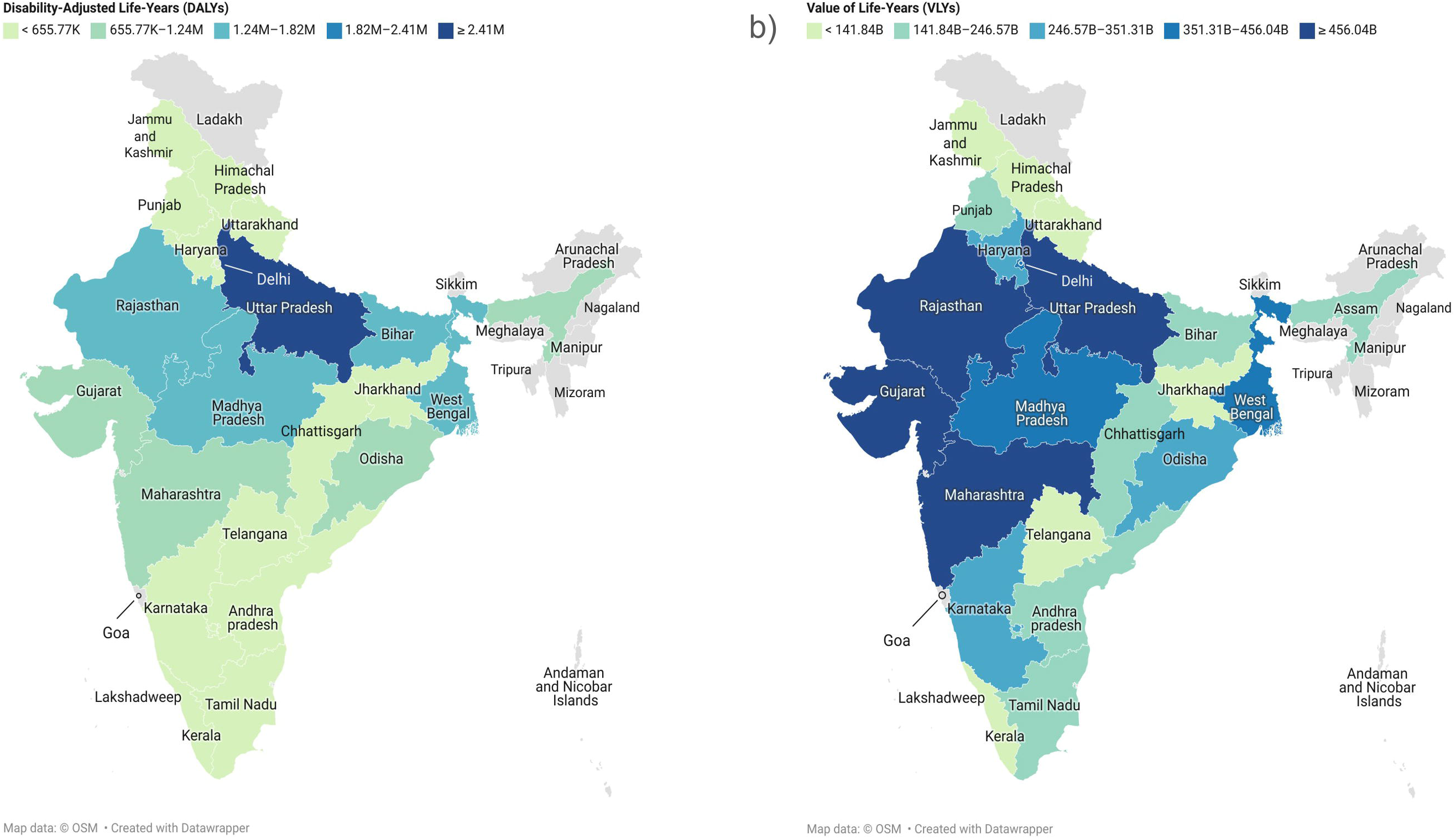
State-wise disease burden of stillbirth for India using the Health Management Information System (HMIS) derived via the YLL approach. **Figure 1b:** State-wise economic burden of stillbirth for India using per capita gross domestic product (GDP).

Other data sources, like the SRS, reported 79,292 stillbirths in 2019, which led to 5.53 million years of life lost. Madhya Pradesh (818,566 life-years) showed the highest disease burden and Telangana (0 life-years) showed the least **(Web Figure 1)**. CRS reported 166,019 stillbirths in 2019, which led to 11.57 million life years lost. The burden ranged from Uttar Pradesh (2.41 million life-years) to Himachal Pradesh (20,468 life-years) **(Web Figure 2)**.

According to sensitivity (Kant) analysis, nationally, stillbirths led to 18.13 million life years lost per HMIS data, 11.49 million life years lost per CRS data, and 5.51 million life years lost per SRS data. We depicted the DALY values for states/UTs in **Web Annexure 1**.

Nationally, there was a 1.22% difference in DALYs from primary and sensitivity analyses. Across states and UTs, the difference ranged from 0.36% (Kerala) to 2.15% (Odisha) **(Table 1)**. Compared to Kant’s approach, the YLL-based approach consistently showed higher burden values, but only marginally. However, we found a significant correlation between the DALYs estimated by the two approaches (r_s_ = 0.99) depicting an agreement between the two approaches across states/UTs.

**Table 1:** Percentage Differences in DALYs obtained by primary and sensitivity analyses.

| **Region** | **DALYs obtained by primary analysis** | **DALYs obtained by sensitivity analysis** | **Percentage Difference** |
| --- | --- | --- | --- |
| Odisha | 970638.8 | 949760.58 | 2.15 |
| Assam | 856710 | 838986.19 | 2.07 |
| Chhattisgarh | 627010.6 | 614698.78 | 1.96 |
| Jammu and Kashmir | 259551.6 | 254798.51 | 1.83 |
| Rajasthan | 1709475 | 1678955.21 | 1.78 |
| Delhi | 379651.8 | 373074.45 | 1.73 |
| Madhya Pradesh | 1572825 | 1546943.66 | 1.64 |
| West Bengal | 1365574 | 1345289.93 | 1.48 |
| Haryana | 467770.8 | 461724.16 | 1.29 |
| Punjab | 358103.2 | 353497.27 | 1.29 |
| Jharkhand | 626335 | 618700.64 | 1.22 |
| Uttarakhand | 129551 | 128008.68 | 1.19 |
| Himachal Pradesh | 72588.3 | 71782.06 | 1.11 |
| Uttar Pradesh | 2988473.6 | 2955405.95 | 1.11 |
| Bihar | 1599212 | 1581920.55 | 1.08 |
| Gujarat | 860371.2 | 851283.84 | 1.05 |
| Karnataka | 617368.5 | 611324.38 | 0.98 |
| Andhra Pradesh | 502504.4 | 497676.07 | 0.96 |
| Maharashtra | 1062437.8 | 1053863.58 | 0.81 |
| Tamil Nadu | 431679.6 | 428973.23 | 0.63 |
| Telangana | 237529.4 | 236260.60 | 0.53 |
| Kerala | 128742.4 | 128269.67 | 0.37 |

Nationally, stillbirths led to an economic burden of INR 6.40 trillion (USD 90.96 billion) in 2019. This was equivalent to 3.9% of India’s 2019 GDP. The economic burden varied from INR 560 billion (Maharashtra) to INR 37 billion (Himachal Pradesh) based on per capita GDP **(Figure 1b)**. Notably, Maharashtra, Uttar Pradesh, Rajasthan, Madhya Pradesh, and West Bengal account for more than 40% of the total economic burden. Himachal Pradesh had the lowest economic burden and Maharashtra had the highest economic burden based on per capita gross, net, and non-health domestic product calculations **(Figure 1b, Web Figures 3-4)**.

## Discussion

In 2019, stillbirths led to 18.3 million years of life lost and a monetary loss of INR 6.4 trillion (USD 90.96 billion) for India. Uttar Pradesh showed the highest disease burden per our primary and sensitivity analyses (see **Web Annexure 1**). Maharashtra contributed to the highest economic burden while Himachal Pradesh led to the lowest. Uttar Pradesh, Maharashtra, Rajasthan, Madhya Pradesh, and West Bengal account for more than 40% of disease and economic burden.

Primarily, we used HMIS data for the calculation of stillbirths associated disease and economic burdens. Previous studies have noted the potential undercounting of stillbirths in SRS estimates. This undercounting was possibly attributed to misclassification of stillbirth under categories such as miscarriages or abortions and related stigma.[15] Hence, we opted not to use SRS and CRS data for primary analysis. Further, HMIS has previously been used for stillbirth rate estimation across districts and is gaining popularity as a source for research and policymaking.[16] Previous review assessing the economic burden of stillbirths in high-income countries (HICs) included four studies from the United Kingdom, the United States, and Australia that used cost-of-illness and other modeling approaches with only one study having mean indirect costs.[4] This study noted the total mean cost per stillbirth as USD 311,944. Our analysis using HMIS data noted 263,342 stillbirths in 2019, bringing the per-stillbirth value to USD 345,406. Hence, it is important to go beyond the direct costing approaches to comprehensively understand the economic burden of stillbirth from a societal perspective. The high burden of stillbirth can be partially attributed to a lack of recognition at policy and community levels.[17] Tracking disease and economic burden along with rates can highlight the true magnitude of the problem.

To our knowledge, no previous studies have looked at the disease burden of stillbirths in DALY terms for India. We present state-level estimates that depict within-country variations. Our analysis also presents burden estimates across data sources. We calculated the economic burden in VLY terms which allowed for comprehensive estimation compared to the traditional human capital approach.

The data for life expectancy for live births was derived from SRS, which only included data for 21 states.[10] Hence, we were unable to include 15 states in the subnational analysis. Our primary estimates are based on HMIS and hence subject to data completeness, coverage, and other quality issues faced by the data source.[18]

## Conclusion

Analyzing subnational variations and incorporating stillbirth burden metrics in child health policies such as INAP could improve resource allocation toward the problem. Measuring and tracking stillbirth disease and economic burdens would represent the true impact of stillbirths on society and help improve investments toward stillbirth reduction as a global public health issue.

## Supporting information

Web Annexure 1, Web Figure 1, Web Figure 2, Web Figure 3, Web Figure 4, Web Figure 5, Web Figure 6, Web Figure 7

## Data Availability

Not available online yet

## KEY MESSAGE

India has the highest absolute number of stillbirths, yet no previous studies have looked at the disease burden of stillbirth at the national or global level. This study looks at the stillbirth disease burden in terms of disability-adjusted life-years (DALYs) and economic burden in value of life-years (VLYs). In 2019, the disease burden was 18.3 million DALYs, and the economic burden of stillbirth was INR 6.4 trillion for India. Analyzing interstate variation and incorporating stillbirth burden metrics in child health policies such as the India Newborn Action Plan could improve resource allocation toward stillbirth.

