## Supplementary material for "Disease and Economic Burden of Stillbirths in India in 2019": Web Annexure 1, Web Figure 1, Web Figure 2, Web Figure 3, Web Figure 4, Web Figure 5, Web Figure 6, Web Figure 7

**SUPPLEMENTAL METHODS**

We calculated the disease burden in terms of disability adjusted life-years (DALYs) using Kant’s approach as our sensitivity analysis.

We first calculated two different stillbirth rates: the still live birth rate (SLBR) using live births (LB) as the denominator **(Supplementary** **Equation** **1)** and the still total birth rate (STBR) using total births, i.e., the sum of stillbirth and live birth as the denominator **(Supplementary Equation 2)**.

$$Still Live Birth Rate \left( SLBR \right)= \frac{stillbirths}{live births}*1000 (Sup Eq 1)$$

$$Still Total Birth Rate \left( STBR \right)= \frac{stillbirths}{total births}*1000 (Sup Eq 2)$$

Typically, life expectancy calculations are based on only live births. Hence, Kant proposed that just as SLBR and STBR are different entities, life expectancies (LE) calculated using live and total births are different entities too - life expectancy of live births (LELB) and life expectancy of total births (LETB). LETB is stillbirth-adjusted life expectancy (SALE) (see **Supplementary Equation 3)**.

$$Stillbirth Adjusted Life Expectancy= \frac{life expectancy of live births \left( LELB \right)*1000}{1000+still live birth rate (SLBR)}(Sup Eq 3)$$

As previously stated, for stillbirths, DALYs equate to years of life lost (YLLs). YLLs are calculated as a product of stillbirths and life expectancy. Since the life expectancy of stillbirths is zero, Kant suggested DALYs be calculated via a different approach. He proposed that stillbirth DALYs be calculated as the absolute difference in life expectancy of live births (LELB) and stillbirth-adjusted life expectancy (SALE) multiplied by the number of live births **(Supplementary Equation 4)**.

$$DALY for SB=YLL for SB=\left| LELB-SALE \right|*LB (Sup Eq 4)$$

Similar to the primary analysis, we extracted values for live births and stillbirths from the Health Management Information System (HMIS) and life expectancy data from the sample registration system (SRS) for India and its 21 states and UTs for the financial year 2019-20.[[2,10](https://sciwheel.com/work/citation?ids=14940080&pre=&suf=&sa=0)] For comparison across data sources, we calculated DALYs using stillbirths derived from SRS (using still total birth rate data) and civil registration system (CRS) for India and its 21 states/UTs.[[10,1](https://sciwheel.com/work/citation?ids=14939712,14939719&pre=&pre=&suf=&suf=&sa=0,0)1]

**SUPPLEMENTAL RESULTS**

The SLBR and STBR for India were 12.4 and 12.25, respectively. Chandigarh had the highest registered SLBR (22.46) and STBR (21.96), while Kerala had the lowest SLBR (3.68) and STBR (3.67). On adjusting, stillbirths decreased the national life expectancy by 0.85 years. The impact of stillbirths on LE was highest in Odisha (1.5 years reduction) and lowest in Kerala (0.27 years reduction).

Nationally, stillbirth led to 18.1 million life-years lost in 2019. Uttar Pradesh (2.9 million life-years) had the highest while Himachal Pradesh (70,000 life-years) had the least disease burden via the Kant approach **(Web Figure 5)** Using other data sources like the SRS, we found that nationally stillbirths led to 5.51 million years of life lost. Madhya Pradesh (813,655 life-years) showed the highest while Telangana (0 life-years) showed the least burden **(Web Figure 6)**. CRS values showed stillbirths led to 11.5 million years of life lost. Uttar Pradesh (2.38 million life-years) showed the highest while Himachal Pradesh (20,407 life-years) showed the least burden **(Web Figure 7)**.

**WEB FIGURES**

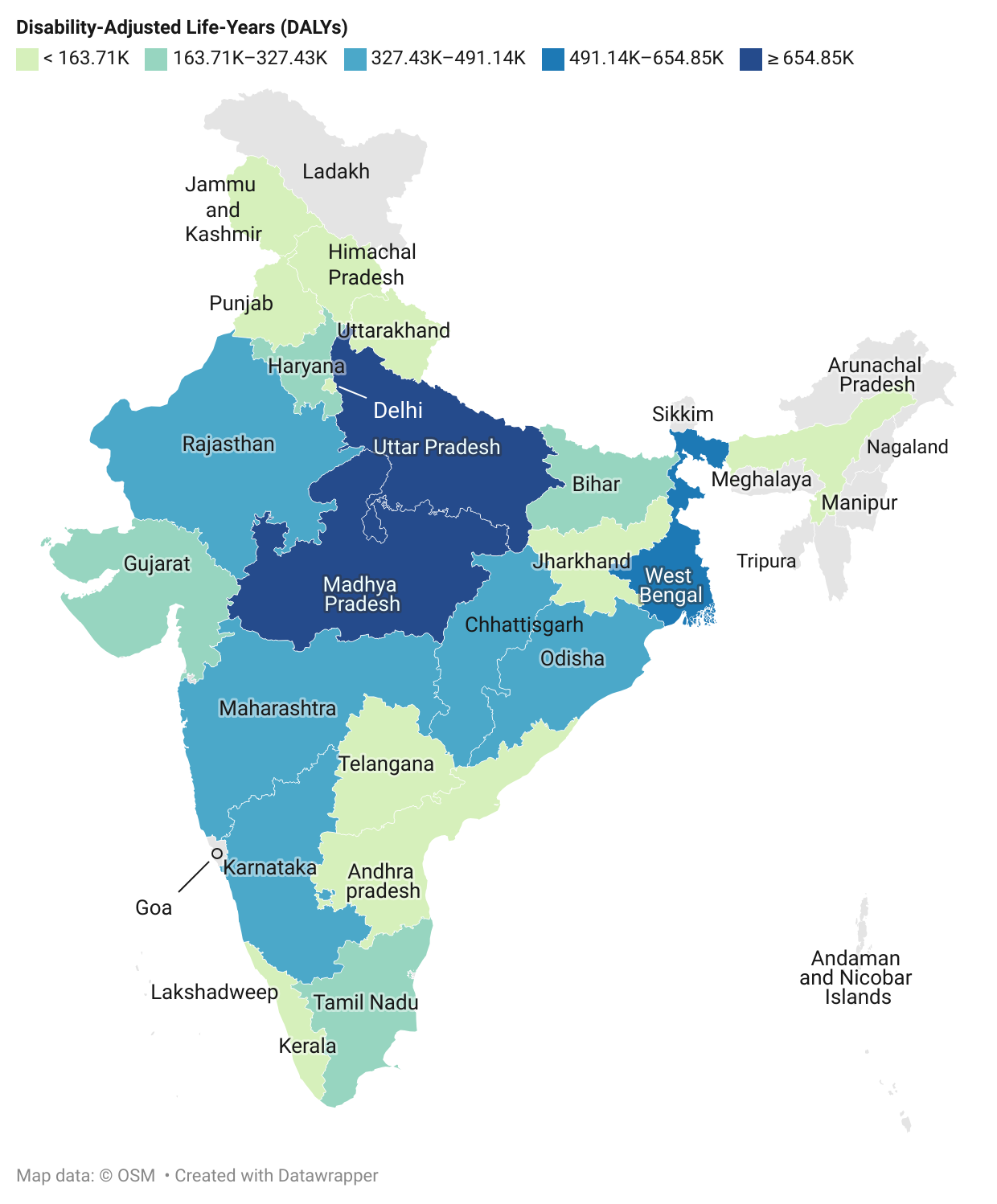

**Web Figure** **1**: State-wise disease burden of stillbirths for India using the Sample Registration System (SRS) derived via the YLL approach.

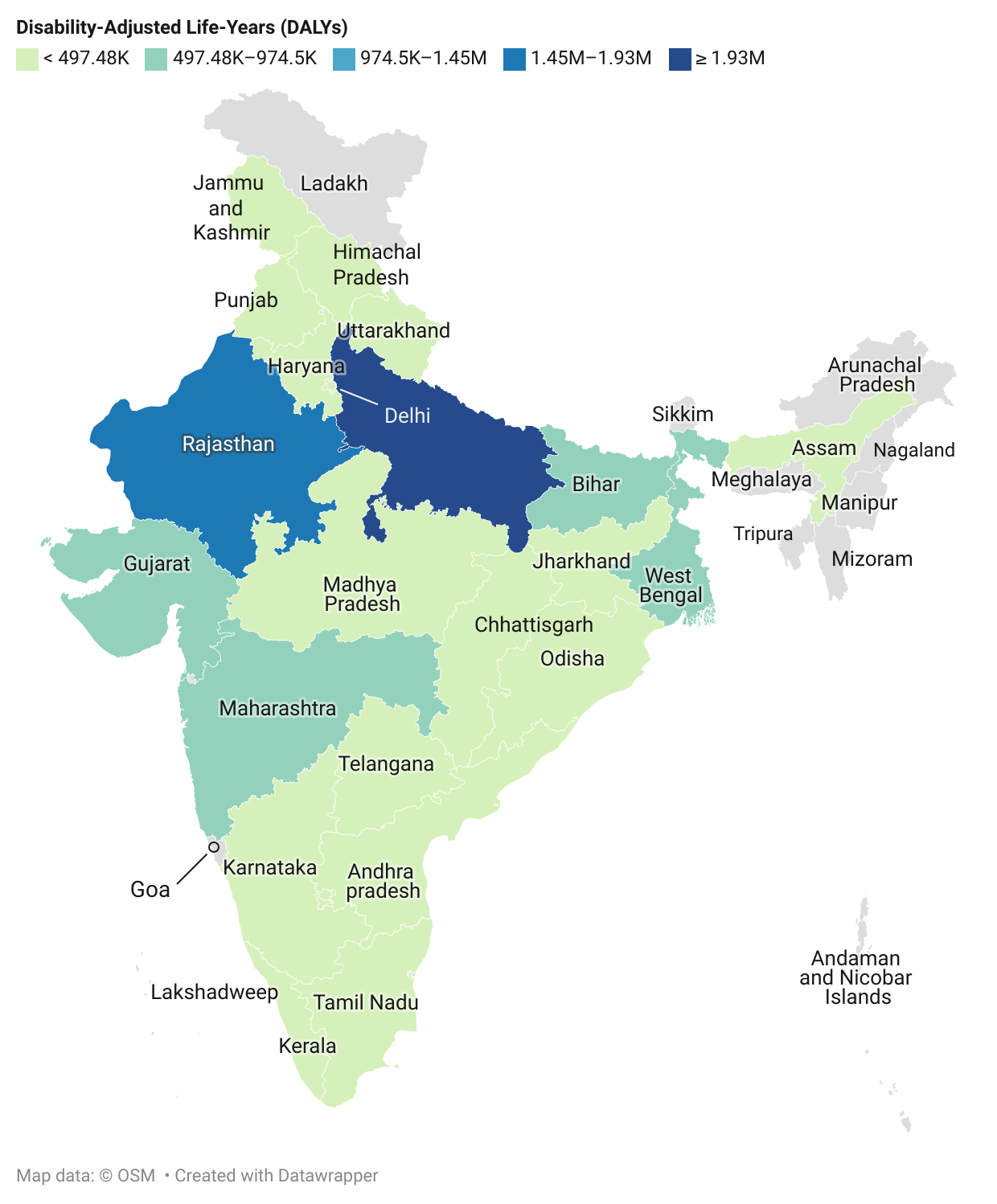

**Web Figure 2:** State-wise disease burden of stillbirth for India using the Civil Registration System (CRS) derived via the YLL approach.

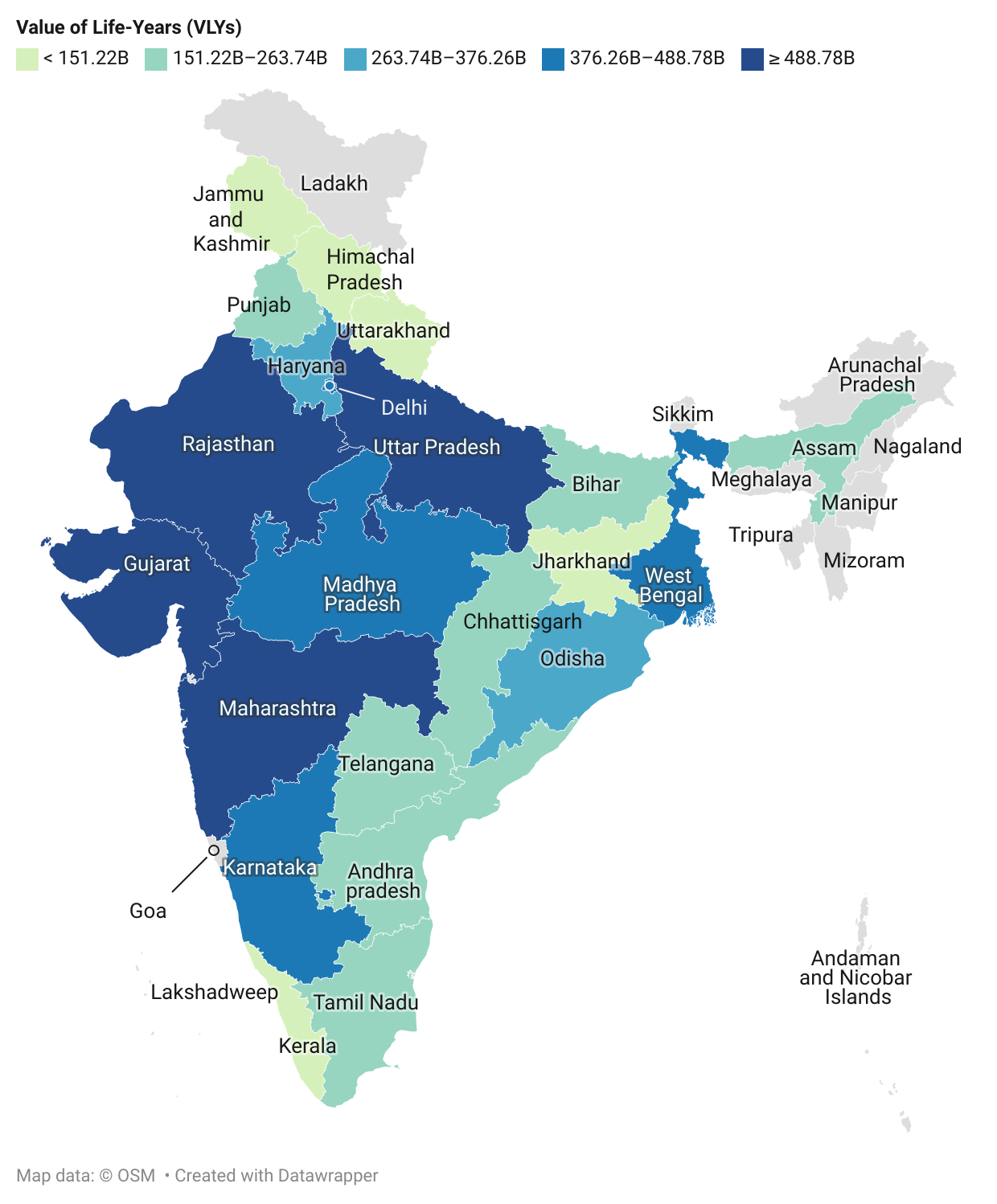

**Web Figure 3**: State-wise economic burden of stillbirth for India using per capita net domestic product (NDP).

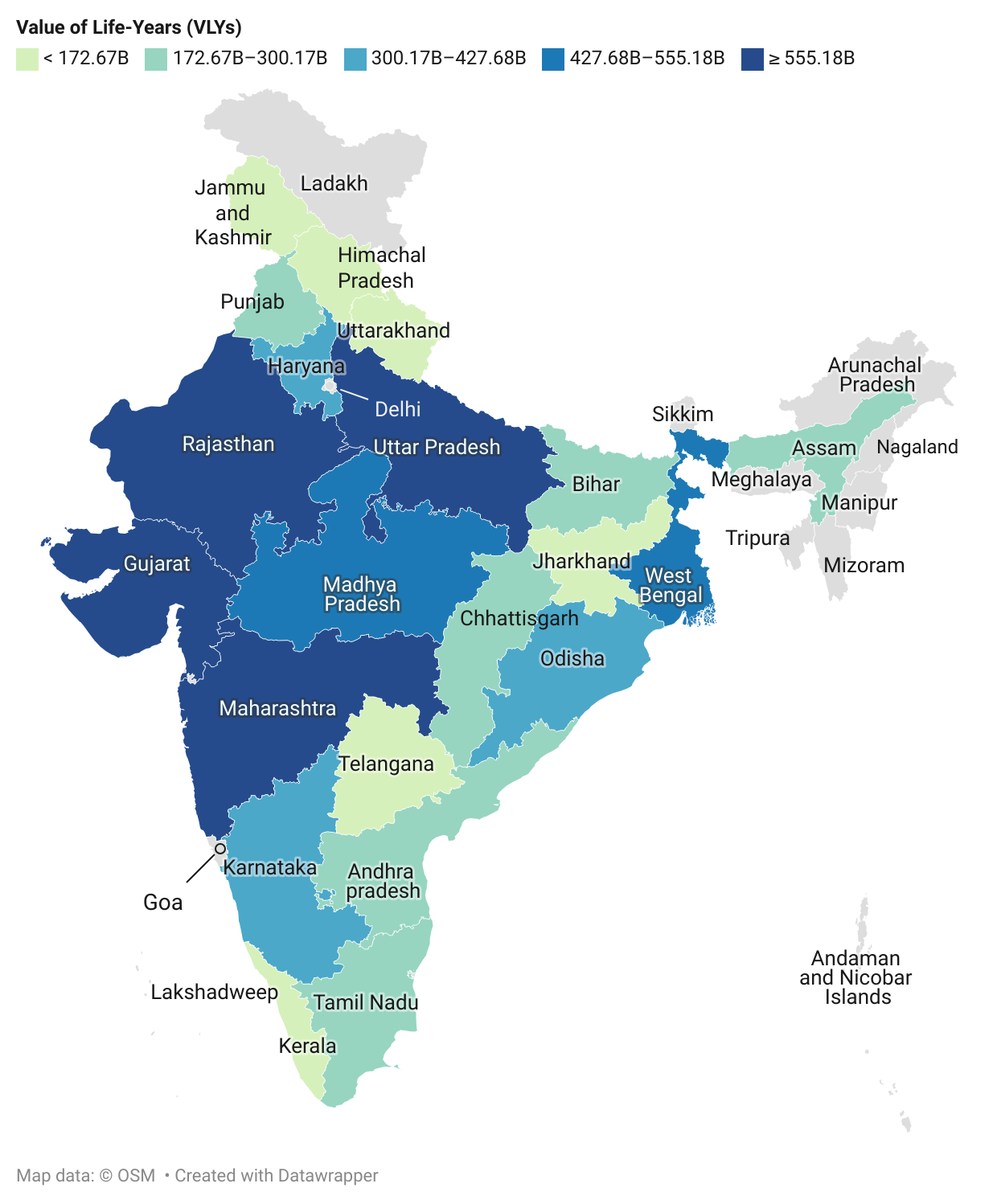

**Web Figure 4:** State-wise economic burden of stillbirth for India using per capita non-health domestic product (NHGDP).

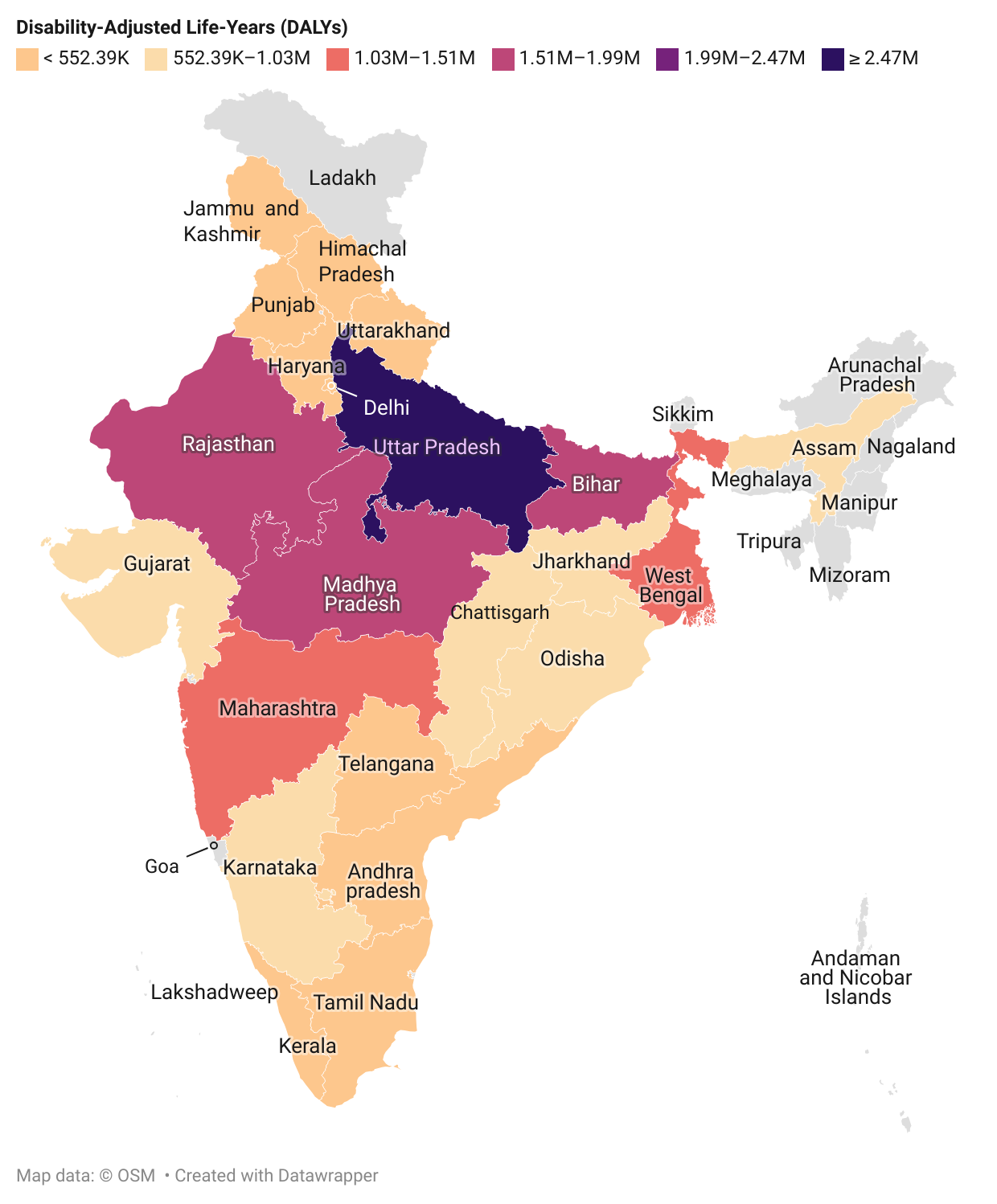

**Web Figure 5:** State-wise disease burden of stillbirths for India using the Health Management Information System (HMIS) derived via the Kant approach.

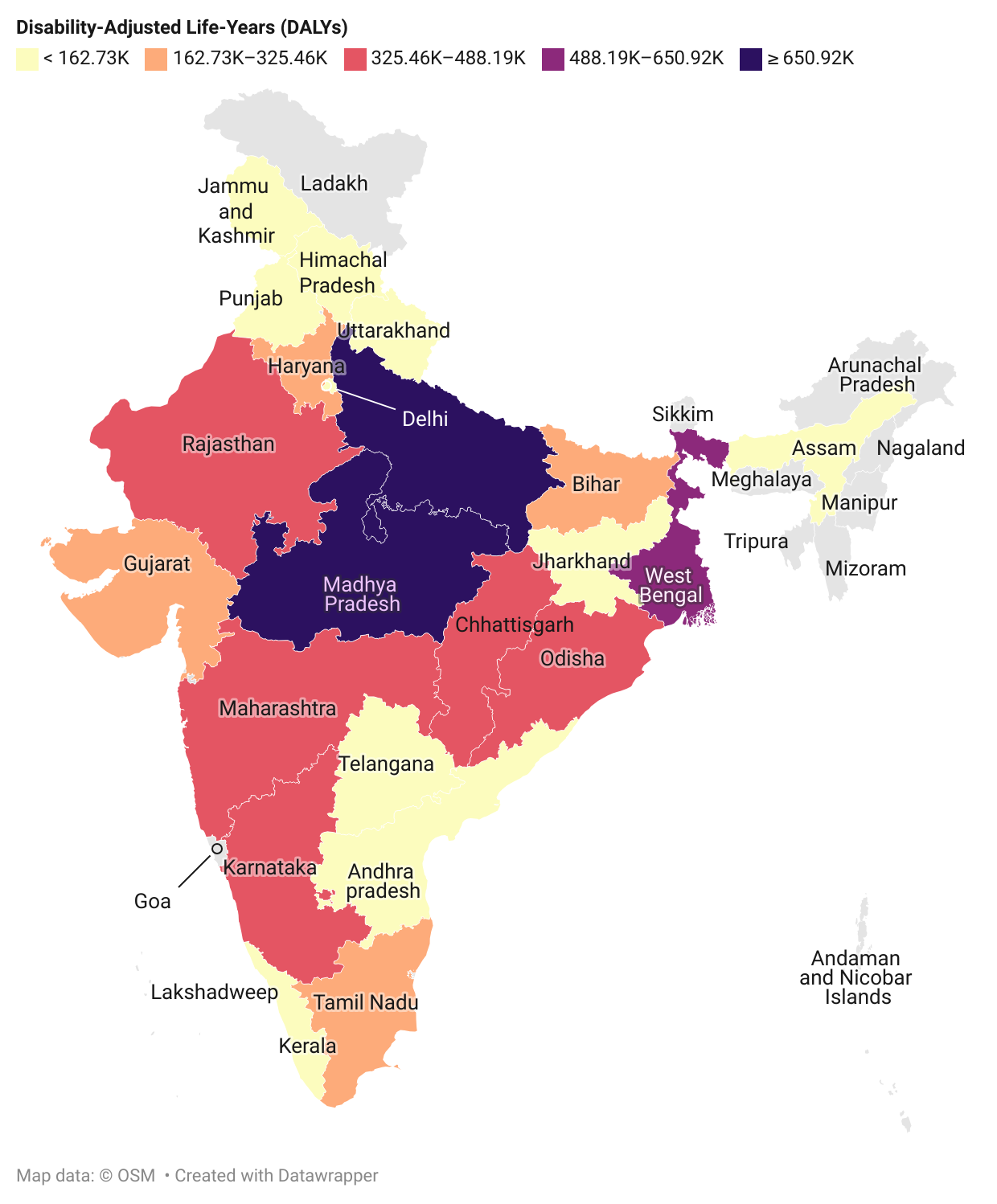
**Web Figure 6:** State-wise disease burden of stillbirths for India using the Sample Registration System (SRS) derived via the Kant approach.

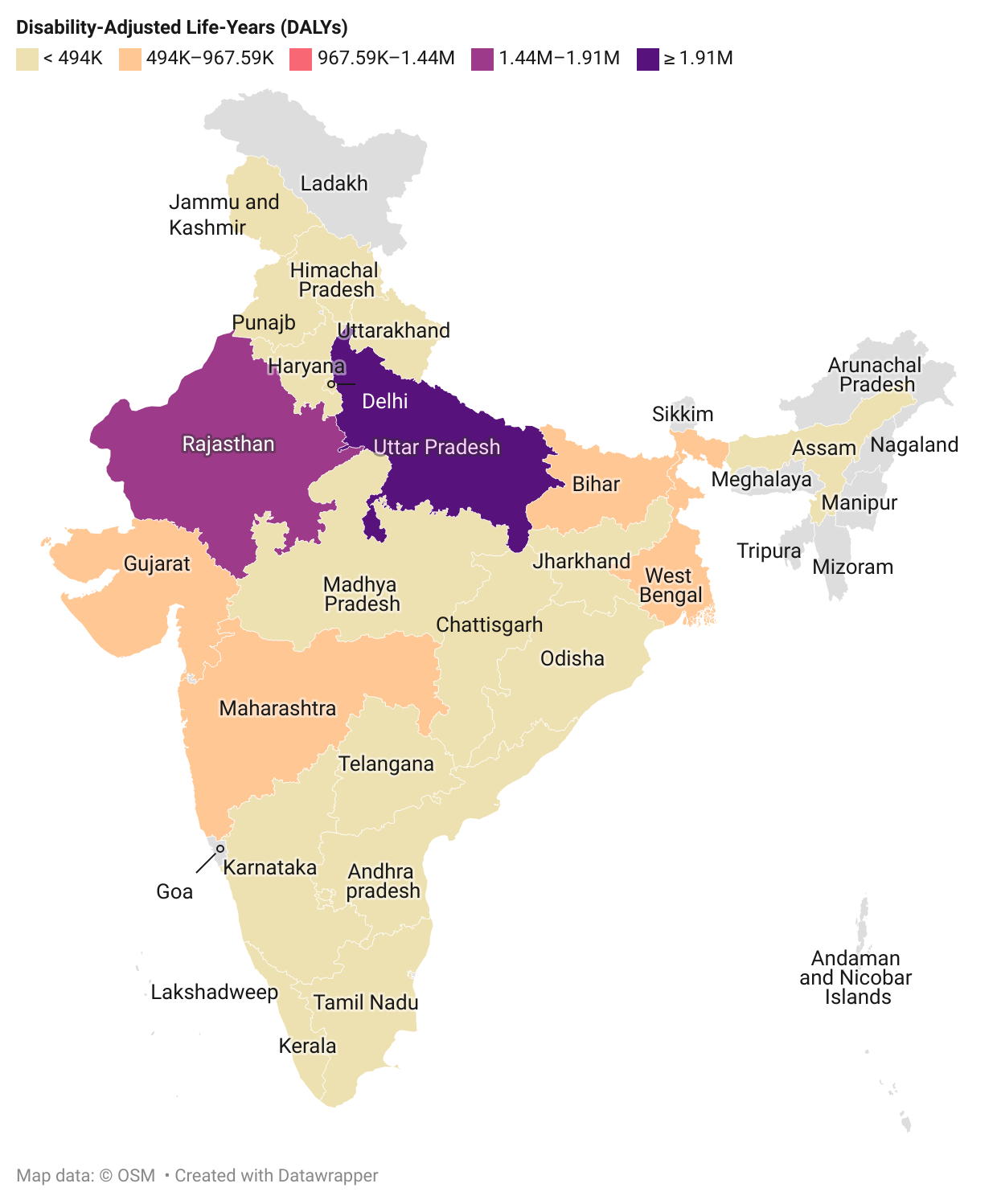

**Web Figure 7:** State-wise disease burden of stillbirth for India using the Civil Registration System (CRS) derived via the Kant approach.
